# Circulating B Cells in Relapsing-Remitting Multiple Sclerosis Show Markedly Different Patterns of Regulatory Marker Expression Compared with Healthy Controls

**DOI:** 10.1101/2022.08.30.22279393

**Authors:** Daniel W. Mielcarz, Alan J. Bergeron, John K. DeLong, Alexandra Dias, Kathleen M. Smith, Karen L. Mack, Lloyd H. Kasper, Jacqueline Y. Channon

**Affiliations:** Departments of Microbiology/Immunology and the Dartmouuth Cancer Center, Geisel School of Medicine at Dartmouth, Lebanon, New Hampshire, USA

## Abstract

Recent evidence has shown that B cells may play a key role in the pathogenesis of relapsing-remitting multiple sclerosis. Studies in our laboratory have shown a difference in the production of IL-6 and IL-10 by B cells isolated from RRMS patients compared with healthy controls. In order to further characterize the nature of the B cells in RRMS patients, we analyzed samples from patients on no disease modifying treatment for B cell expression of multiple phenotypic and regulatory markers. We observed an increased frequency in the number of circulating B cells, an increase in B1 B cells and a decrease in memory B cells in RRMS patients. These B1 cells showed a significantly higher frequency of CD5 expression and the memory B cells a significant increase in the class-switched IgD-phenotype. We also examined death receptors involved in apoptotic pathways. CD95 frequency was significantly lower in RRMS patients tan healthy controls in all B cell subsets. Conversely, frequency of PD-1 was elevated in both the naïve and memory B cell subsets, and PD-L1 was elevated in B1 cells from RRMS patients. Finally, we examined a series of immunoreceptor tyrosine-based inhibition motif (ITIM)-containing inhibitory receptors, including members of the SIGLEC family. Significantly higher levels of CD22, CD305 and CD307d were seen in RRMS patients, while significantly lower levels of SIGLEC-10 were observed. Taken together, these results indicate a potential for differential regulation of B cells in RRMS patients that may provide an avenue for B cell directed therapies for the disease.

## Introduction

Multiple sclerosis (MS) is a chronic inflammatory disease of the CNS leading to demyelination and neurodegeneration, and relapsing-remitting multiple sclerosis (RRMS) is associated with acute inflammatory episodes resulting in a reduction in neurological function separated by periods of remission^1^. T_h_1 and T_h_17 subsets of myelin-specific CD4+ cells have traditionally been thought to be the main pathogenic cells in RRMS^2^. However, recent studies provide strong evidence for a role for B cells in the pathology of RRMS. It is known that CSF samples from patients with active RRMS are enriched in memory B cells and that persisting plasmablasts are a major source of antibody-secreting cells responsible for oligoclonal bands in the CSF^3^. In addition to oligoclonal bands found in the CSF, MS patients have also been shown to have a unique autoantibody signature in their serum^4^. Therapy directed at the depletion of CD20+ B cells has been shown in Phase II clinical trials to be effective in reducing relapse rate and reducing lesions as detected by MRI in those with RRMS^5, 6^. Interestingly, a Phase II trial with atacicept, a soluble inhibitor of B cell survival, showed higher levels of inflammation and MRI activity in treated patients, indicating that there may be a pleiotropic effect of B cells in RRMS^7^. These two studies and others suggest a role for B cell regulation and homeostasis in the pathogenesis of RRMS.

B cell homeostasis has been implicated in several autoimmune diseases, including RRMS. One means by which the immune system controls immune responses is through Fas/FasL interactions. In general, binding of Fas(CD95) by FasL leads to apoptosis of the cell expressing Fas^8^. This mechanism typically occurs as an immune response is ending, as a way to prevent runaway inflammation. Fas and FasL have been shown to be present at high levels in brain lesions of MS patients^9^. Examination of Fas/FasL regulation in RRMS, has focused on T cells, however studies from other disorders and animal models shed light on important roles for Fas in B cell development, regulation and homeostasis. Ligation of Fas on B cells has been specifically shown to lead to apoptosis of B cells in vitro ^10^. Lack of Fas expression in germinal center B cells has been shown to lead to aberrant proliferation of both T and B cells and defects in negative selection against autoantigens ^11^.

Another way in which the immune system regulates itself is through PD-1/PD-L1/PD-L2 interactions. PD-1 (programmed cell death-1) - a member of the CD28/CTLA4 family of receptors - is expressed on activated lymphocytes and serves as a checkpoint by which active immune responses are controlled and ended following pathogen clearance to prevent ongoing inflammation and autoimmunity^12, 13^. PD-L1 and PD-L2 are the ligands for PD-1 whose interactions lead to a down regulation of immune response and/or apoptosis in the cell expressing PD-1. PD-L1 is expressed on a broad range of tissues, while PD-L2 is restricted to cells of hematopoetic origin^14^. The involvement of PD-1 in autoimmunity is supported by the fact that PD-1 knockout mice develop a spontaneous autoimmune disease resembling human lupus or autoimmune cardiomyopathy depending on genetic background^15, 16^. Furthermore, in EAE, blockade of PD-1 or PD-L1 exacerbates disease^17^. In MS, these markers have been shown to be modulated depending on whether the subject is exhibiting active or stable disease, with a reduction of PD-1 on T cells and a reduction of PD-L1 on monocytes in active compared with stable disease, potentially indicating an important role for this receptor ligand pair in RRMS^18^.

Immunoreceptor tyrosine-based inhibition motif (ITIM) is a protein domain present on many transmembrane proteins found on immune cells that downregulates activation^19^. The ITIM domain becomes phosphorylated upon ligation, and it in turn activates phosphatases that downregulate activation signal transduction^19^. Prominent members of the ITIM-containing family of receptors are the sialic acid-binding immunoglobulin-type lectins or siglecs. These siglecs are often involved in the downregulation of immune responses. Two siglecs of particular interest are Siglec-10 and CD22. CD22 is found on mature B cells and downregulates BCR-driven immune responses. Siglec-10 associates with CD24 to downregulate immune responses. Other ITIM-containing proteins of interest are CD305 and CD307d. CD305, or Leukocyte-associated immunoglobulin-like receptor 1 (LAIR-1) is thought to be an inhibitory receptor. It is expressed on B cells and T cells, and specifically has been seen on most naïve and memory B cells ^20^. Mice lacking the LAIR-1 gene show defects in B cell function^21^. CD307d, or Fc receptor like protein 4 (FcRL4) is also an inhibitory receptor but is seen on a smaller fraction of B cells^20^. CD307d expression was noted in HIV-infected individuals on a subset of exhausted tissue-resident memory B cells^22^. Recently, CD307d has been shown to efficiently bind IgA, indicating a role for this inhibitory receptor in biding immune complexes^23^.

Recent studies in our laboratory have shown important differences in IL-6 and IL-10 cytokine production by B cells isolated from RRMS patients compared with healthy controls, as well as changes in B cell subset composition (manuscript in preparation). To further examine differences between RRMS patients and healthy controls, we examined expression of the previously described proteins related to B cell function, regulation and homeostasis via flow cytometry.

## Materials and Methods

### Patients

For this trial, 10 patients not on disease modifying therapy with clinically defined RRMS (McDonald criteria) were enrolled and treated with IFNβ-1b every other day (qOD). The treatment window was 6 months during which time 200 ml blood was drawn and assayed for immunologic changes at baseline, month 2 and month 6 post-therapy. The principal immunologic outcome was the effect of therapy on B cells as determined by phenotypic changes and intracellular cytokine expression using cryopreserved PBMCs. In this study, baseline samples were examined for differences with 10 healthy controls. This study was approved by the Geisel School of Medicine at Dartmouth Institutional Review Board.

### PBMC Isolation and cryopreservation

Blood was collected in 200ml blood bags containing sodium citrate. Peripheral blood mononuclear cells were isolated via density gradient separation. Briefly, 20 ml of blood was overlaid with 20 ml of Histopaque-1077. Blood was centrifuged at 1800 rpm for 30 minutes at 25°C. Following centrifugation, PBMCs were removed and washed 2 times via centrifugation with HBSS without calcium and magnesium ions. PBMCs were counted and adjusted to desired concentration in human AB serum + 10% DMSO. PBMCs were then frozen overnight at -80°C in a cryopreservation chamber, then transferred to -140°C for long-term storage.

### Flow cytometry for B cell subset analysis

A violet amine viability dye (Invitrogen) and anti-CD14-Pacific blue was present in all panels as a dump channel and viability after thawing and washing was >80% of thawed cells. Antibodies were from BD or from BioLegend and were all titrated before use (see Table 1 for antibody source and clone information). Each panel contained duplicate fully-stained samples and a fluorescence-minus-one (FMO) tube for each antibody. Patient PBMCs were thawed on the same day and stained in 96-well plates. Healthy controls (10 samples) were thawed on a different day and stained in the same way as for patient samples. Inter-day staining variability was controlled for by use of an aliquot of cryopreserved PBMCs from the same donor that was thawed and stained daily in parallel with patient or healthy control PBMCs. Flow cytometry was carried out using a 10-color Gallios cytometer (Beckman Coulter). FlowJo software (Tree Star, Inc.) was used for post-hoc compensation and analysis.

### Statistical analysis

Groups were compared using an unpaired, two-tailed t test with Welch’s correction using Prism software (GraphPad).

## Results

### Differing frequencies of B cell subsets are seen in RRMS patients and healthy controls

Circulating mature CD19+/CD20+ B cells can be divided into three main subsets: naïve (CD27-CD43-), memory (CD27+CD43-), and B1 (CD27+CD43+)^24^.Cryopreserved peripheral blood mononuclear cells (PBMC) from RRMS patients and age- and gender-matched healthy controls were thawed and stained for flow cytometry as described in the materials and methods to determine frequencies of these B cell subsets. Subsets were gated as shown in Figure 1A. RRMS patients had a significantly higher (p<0.05) frequency of total B cells (Figure 1B) compared to healthy controls. Examining subset frequencies, RRMS patients had a lower frequency of memory B cells (p<0.05) and a higher frequency of B1 B cells (p<0.01) and a trend toward in an increase in naïve B cells (Figure 1C). In the mouse, B1 cells can be divided into CD5+ and CD5-types (B1a and B1b, respectively), and CD5+ B cells have been shown to be increased in the CSF of MS patients^25, 26^. To that end, we examined CD5 expression on B1 cells from RRMS patients and healthy controls. RRMS patients had a significantly higher (p<0.01) frequency of CD5+ B1 B cells (Figure 1D). Memory B cells can be further subdivided into unswitched (IgM+/IgD+) and class switched (IgM-/IgD-) subsets^27^. RRMS patients had a significantly higher (p<0.05) proportion of switched memory B cells, indicating a more antigen-experienced phenotype (Figure 1E).

**Figure 1.**
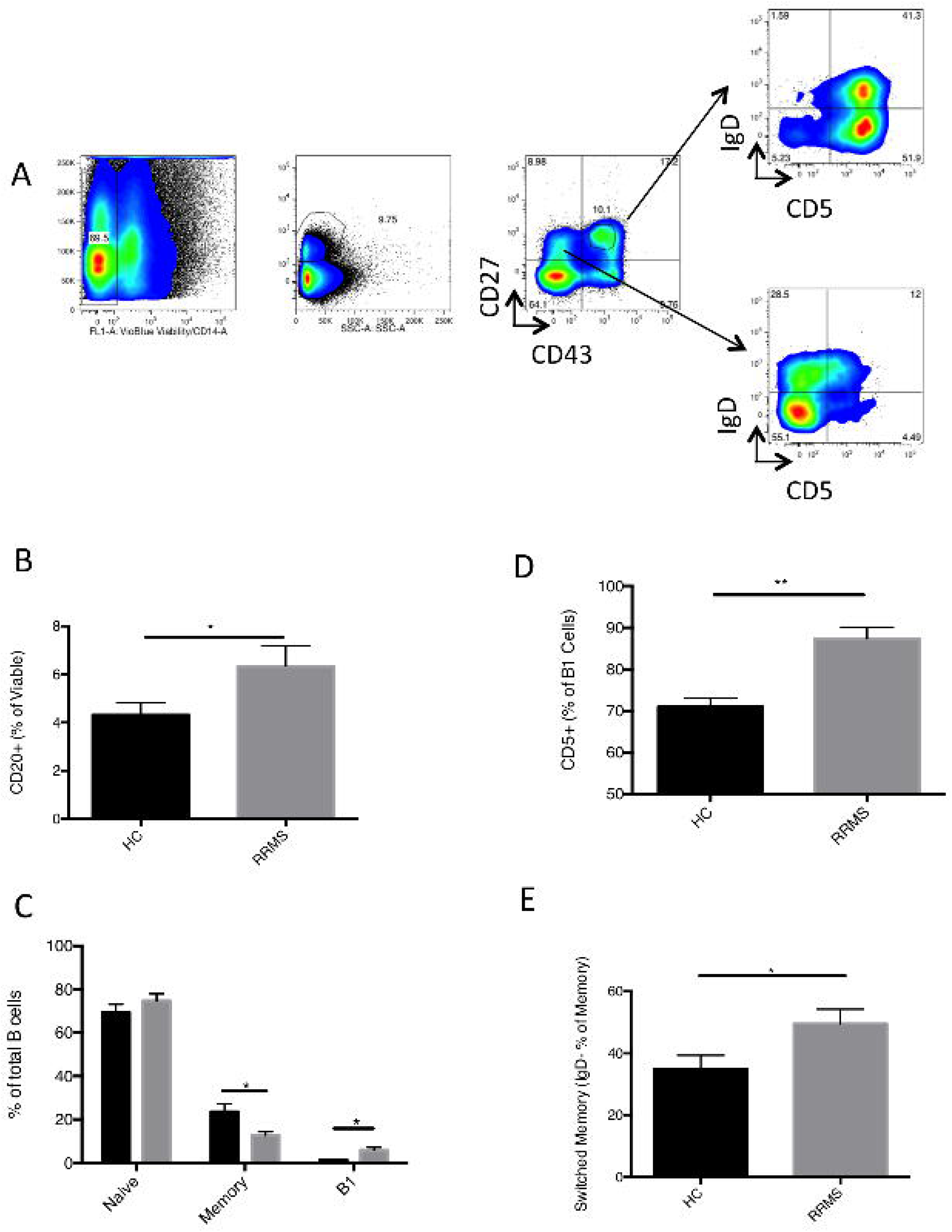
B cell subset frequencies differ in RRMS patients and healthy controls. Cryopreserved and thawed PBMC from 10 RRMS patients and 10 healthy controls were analyzed via flow cytometry. (A) Gating strategy for delineating subsets. From left to right: viable, CD14-cells were gated, then CD20+ B cells. CD27 and CD43 expression was used to delineate naïve B cells (CD27-CD43-), memory B cells (CD27+CD43-) and B1 B cells (CD27+CD43+). Representative data from 1 RRMS patient at baseline is shown. The following graphs represent 10 healthy controls and 10 RRMS patients. Samples were analyzed in triplicate, and error bars represent the standard error of the mean. Groups were compared by unpaired, two-tailed t test with Welch’s correction * p<0.05 ** p<0.01. (B) Total B cell frequencies. (C) B cell subset frequencies. (D) Frequency of CD5 expression on on B1 B cells. (E) Frequency of switched memory B cells.

### Apoptosis inducing and related checkpoint molecules are differentially expressed on B cells from RRMS patients and healthy controls

A key mechanism of immune control involves the induction of apoptosis through death receptors on B and T cells. We examined two of these inhibitory receptors: CD95(Fas) and PD-1. Healthy controls had a consistently high frequency of CD95 on all three subsets of B cells examined, particularly the memory and B1 subsets where the frequency approached 100% (Figure 2A). In contrast, RRMS patients had significantly lower frequencies of CD95+ naïve (p<0.0001), memory (p<0.0001) and B1 B cells (p<0.0001). Interestingly, among the CD95+ cells in the B1 subset, a significant (p<0.0001) reduction in mean fluorescent intensity was observed (Figure 2B). This reduced expression may make FasL-triggered homeostasis more difficult for these B cells, particularly in the B1 subset, perhaps contributing to the pathogenesis of RRMS. PD-1 is a surface molecule, that when ligated, decreases the activation state of the cell expressing it. Unlike the lower levels of CD95 seen, RRMS patients had a significantly higher frequency of PD-1 expressing naïve (p<0.01) and memory (p<0.001) B cells (Figure 3A), compared with the very low levels (∼2-5%) seen in healthy controls. B1 B cells showed a trend toward higher frequency that was not statistically significant (Fig. 3A), but all three B cell subsets showed significant increases in expression of PD-1 as measured by MFI, with B1 B cells expression the highest levels of PD-1 (Fig. 3B). In contrast to the CD95 results, these elevated levels of PD-1 suggest that the B cells from RRMS patients may be more susceptible to regulation, although the frequency of PD-1 positive B cells is much lower than that of the CD95 B cells.

**Figure 2.**
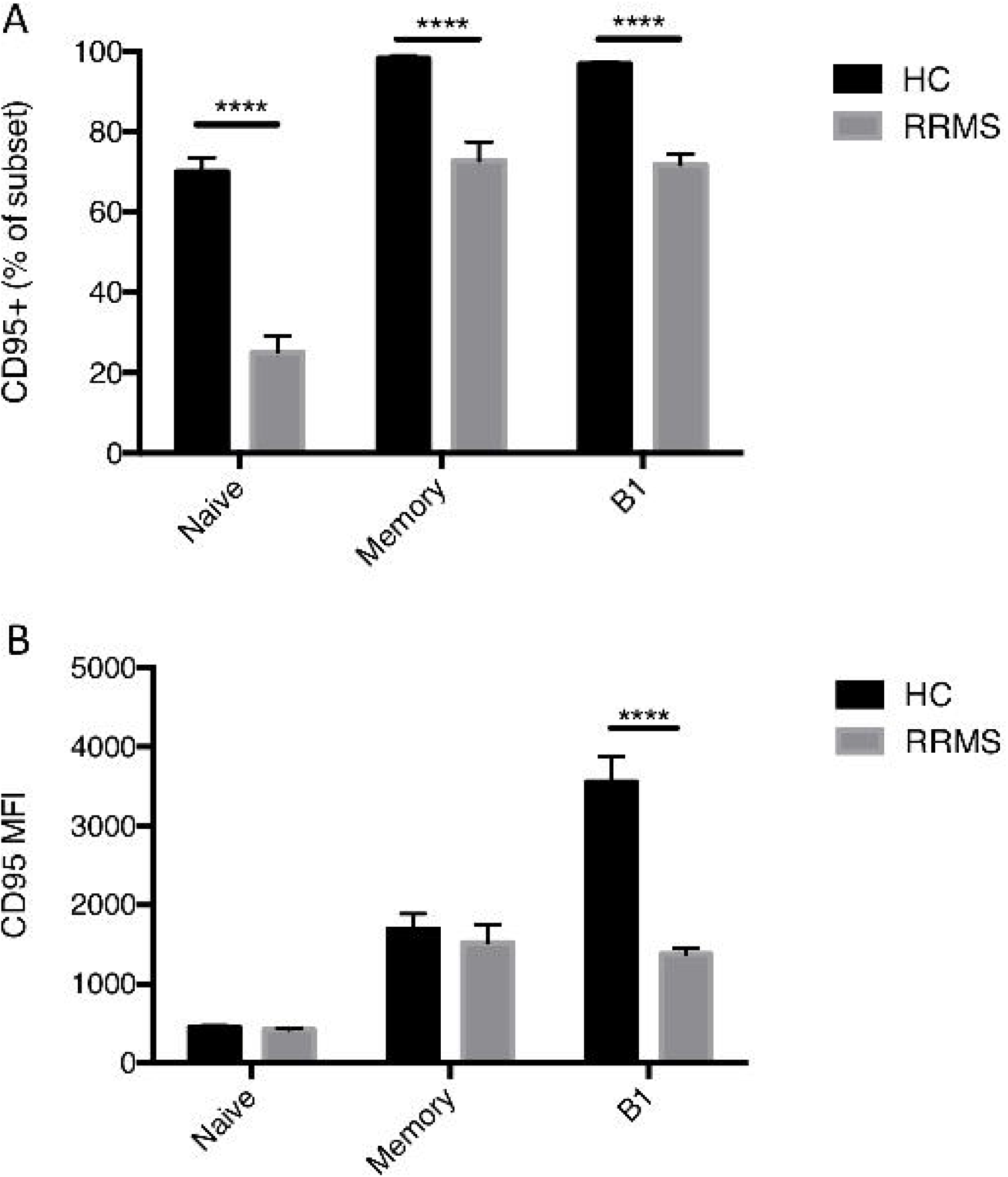
CD95 expression on B cell subsets is higher in healthy controls than RRMS patients. Cryopreserved and thawed PBMC from 10 RRMS patients and 10 healthy controls were analyzed via flow cytometry. Cells were gated as in Figure 1 and examined for CD95 expression. Samples were analyzed in triplicate, and error bars represent the standard error of the mean. Groups were compared by unpaired, two-tailed t test with Welch’s correction **** p<0.0001. (A) CD95 frequency on B cell subsets. (B) CD95 geometric mean fluorescence intensity (MFI) on B cell subsets.

**Figure 3.**
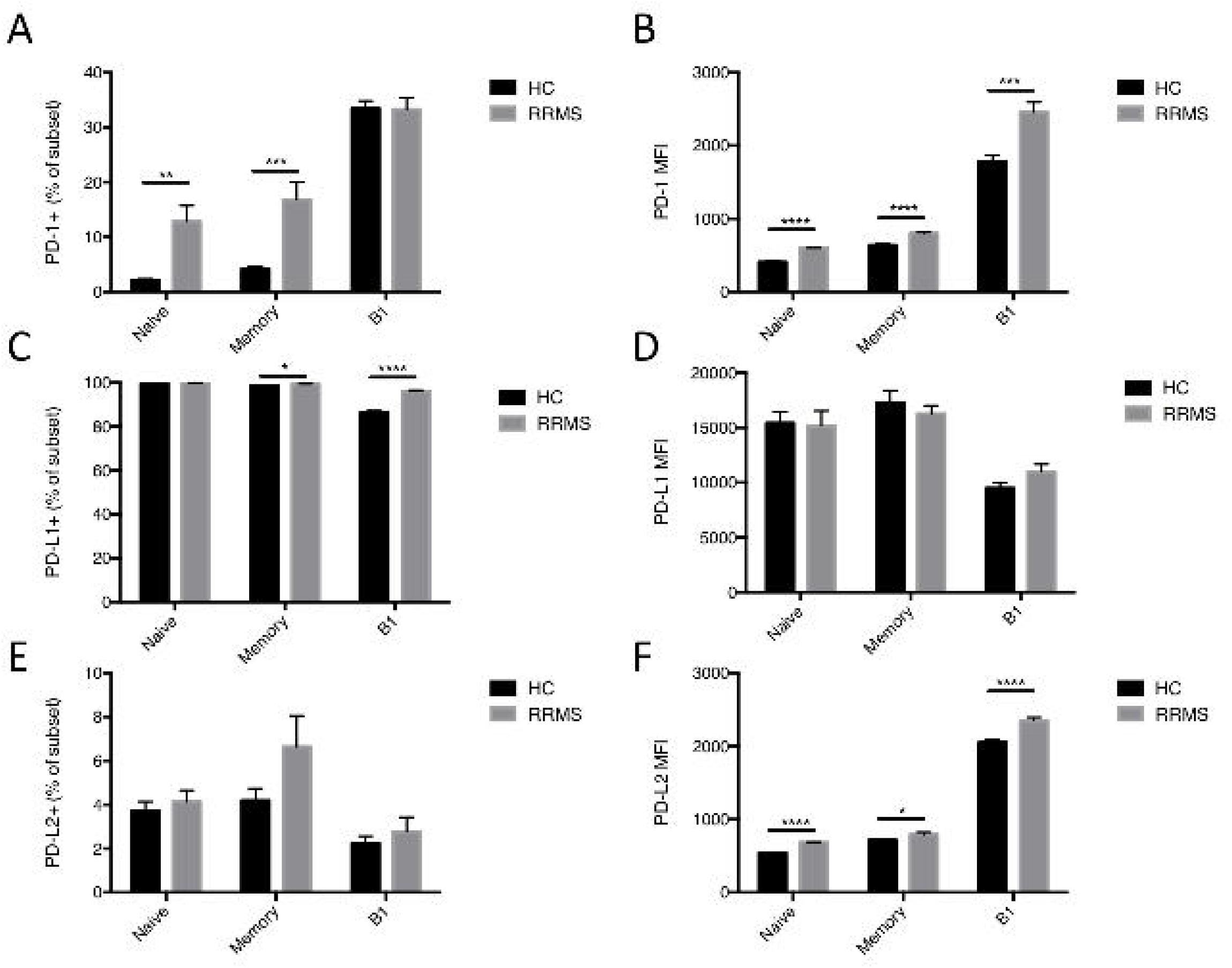
PD-1, PD-L1 and PD-L2 expression on B cell subsets differs in healthy controls and RRMS patients. Cryopreserved and thawed PBMC from 10 RRMS patients and 10 healthy controls were analyzed via flow cytometry. Cells were gated as in Figure 1 and examined for PD-1, PD-L1 and PD-L2 expression. Samples were analyzed in triplicate, and error bars represent the standard error of the mean. Groups were compared by unpaired, two-tailed t test with Welch’s correction * p<0.05, ** p<0.01 *** p<0.001 **** p<0.0001. (A) PD-1 frequency on B cell subsets. (B) PD-1 geometric mean fluorescence intensity (MFI) on B cell subsets. (C) PD-L1 frequency on B cell subsets. (D) PD-L1 geometric mean fluorescence intensity (MFI) on B cell subsets. (E) PD-L2 frequency on B cell subsets. (F) PD-L2 geometric mean fluorescence intensity (MFI) on B cell subsets.

In addition to CD95 and PD-1, the frequency of expression of PD-L1 and PD-L2 – the apoptosis inducing ligands of PD-1 – on B cells was also examined. PD-L1 showed constitutive expression (near 100%) on both naïve and memory B cells in both controls and RRMS patients (Fig. 3C). However, while B1 B cells from RRMS patients had a nearly constitutive level of PD-L1 expression, B1 B cells from healthy controls had a mean expression of 87%, a statistically significant lower level (Figure 3C; p<0.001). While no differences were seen between HCs and RRMS patients, PD-L1 was expressed at a lower level on the B1 B cell subset compared with the naïve and memory subsets (Figure 3D). This differential expression in B1 B cells may indicate an increased ability for this population of B cells in RRMS patients to reduce activation in PD-1 expressing T cells and B cells. PD-L2 was expressed on a very low frequency of B cells in both healthy controls and RRMS patients, and at similar levels (Fig. 3E), but the PD-L2+ fraction from all subsets showed a significant increase in expression level as measured by MFI (Figure 3F; p<0.0001 for naïve and B1; p<0.05 for memory). This increased level of may indicate an increase ability for the three B cell subsets to inhibit activation of PD-1 expressing cells.

### Immunoreceptor tyrosine-based inhibition motif (ITIM)-containing inhibitory receptors show differing levels of expression in RRMS and healthy controls

Sialic acid-binding immunoglobulin-type lectins - Siglecs – are a family of lectins chiefly found on immune cells. The majority of siglecs contain an ITIM that, when phosphorylated, inhibits immune activation. We examined two siglecs known to be present on B cells: CD22 and siglec-10. CD22 is phosphorylated upon B cell receptor activation and activates a signaling pathway that regulates BCR-mediated cellular activation through Ca+ modulation. In naïve and memory B cells, CD22 frequency is present at constitutive levels in both RRMS patients and healthy controls (data not shown). However, in B1 B cells, RRMS patients show a significantly lower frequency (p<0.001) of CD22 expression, with a mean frequency of 76.5% compared to healthy controls at 94.5%. This suggests that B1 B cells from RRMS patients may be less able to control BCR-mediated activation. Like CD22, Siglec-10 is an ITIM containing transmembrane protein found on B cells. Siglec-10 shows homology to CD33 and associates with CD24 to act as an inhibitor of tissue-damage-associated immune responses. In all three B cell subsets examined, we see a significant reduction in frequency of siglec-10 expression (Figure 4B; naïve B, p<0.01; memory B, p<0.05; B1 B, p<0.0001). As with CD22, the most dramatic reduction is seen in the B1 B cell subset, with a mean expression in healthy controls of 60% reduced to 20% in RRMS patients. Taken together, the evidence shows that B cells from RRMS patients, particularly the B1 B cell subset, will be less able to control immune activation through ITIM-containing siglecs.

**Figure 4.**
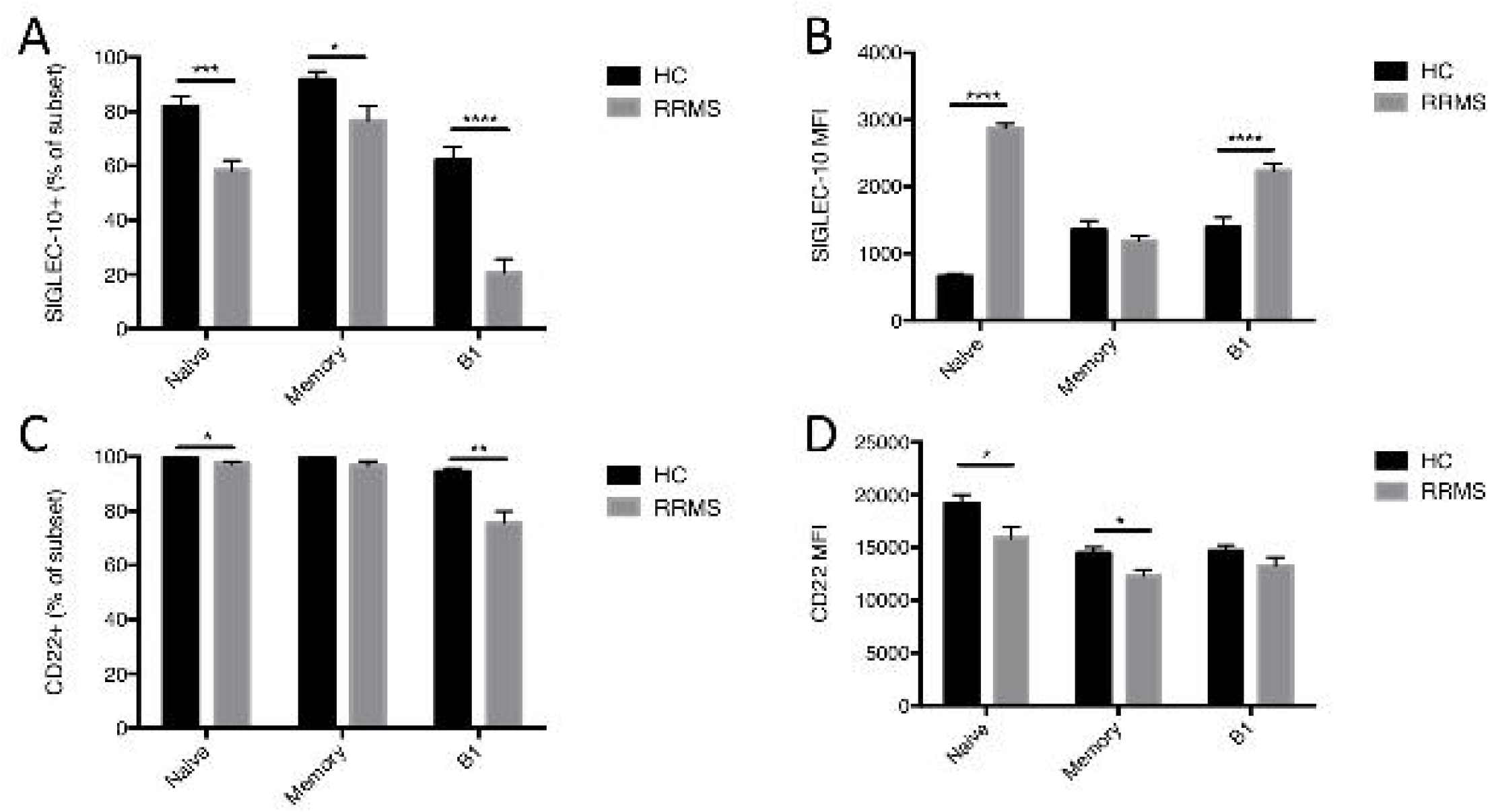
SIGLEC expression on B cell subsets shows marked differences in healthy controls and RRMS patients. Cryopreserved and thawed PBMC from 10 RRMS patients and 10 healthy controls were analyzed via flow cytometry. Cells were gated as in Figure 1 and examined for SIGLEC-10 and CD22 expression. Samples were analyzed in triplicate, and error bars represent the standard error of the mean. Groups were compared by unpaired, two-tailed t test with Welch’s correction * p<0.05, ** p<0.01 *** p<0.001 **** p<0.0001. (A) SIGLEC-10 frequency on B cell subsets. SIGLEC-10 geometric mean fluorescence intensity (MFI) on B cell subsets. CD22 frequency on B cell subsets. (D) CD22 geometric mean fluorescence intensity (MFI) on B cell subsets.

To gain a fuller understanding of ITIM-mediated control of B cell activation, expression of CD305 (LAIR1) and CD307d (FcRL4) - which contain ITIM motifs - were examined. Similar to the siglecs, CD305 and CD307d act to inhibit activation in immune cells. However, where the siglecs examined showed lower levels of expression in RRMS patients, both CD305 and CD307d showed higher levels of expression. The frequency of CD305 in both memory B (p<0.05) and B1 B (p<0.0001) was increased in RRMS patients compared with healthy controls (Figure 5A). In healthy individuals, CD307d+ B cells are normally restricted to the mucosa and mesenteric lymph nodes, with low frequencies found in circulation. We observed a similar result, however, in RRMS patients we observed a statistically significant increase in CD307d+ cell frequency in both the naïve (p<0.0001) and memory (p<0.0001) B cell subsets. Given that both of these markers (CD305 and CD307d) are inhibitory, it may be that this increase seen in RRMS is a compensatory mechanism to deal with lower levels of the ITIM-containing siglecs.

**Figure 5.**
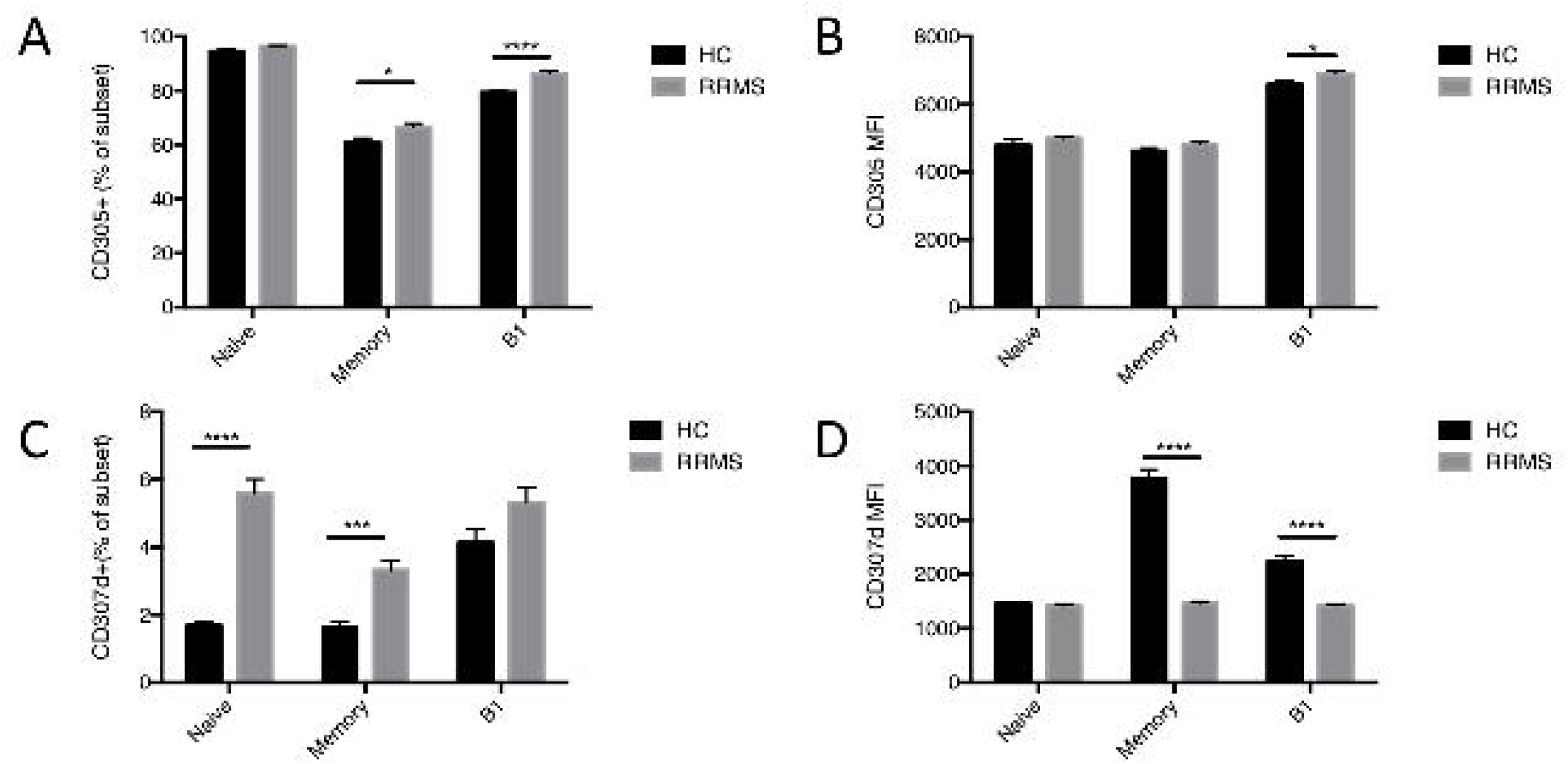
Healthy control and RRMS patients have differing levels of expression the ITIM-containing markers CD305 and CD307d. Cryopreserved and thawed PBMC from 10 RRMS patients and 10 healthy controls were analyzed via flow cytometry. Cells were gated as in Figure 1 and examined for CD305 and CD307d expression. Samples were analyzed in triplicate, and error bars represent the standard error of the mean. Groups were compared by unpaired, two-tailed t test with Welch’s correction * p<0.05, ** p<0.01 *** p<0.001 **** p<0.0001. (A) CD305 frequency on B cell subsets. (B) CD305 geometric mean fluorescence intensity (MFI) on B cell subsets. (C) CD307d frequency on B cell subsets. (D) CD307d geometric mean fluorescence intensity (MFI) on B cell subsets.

## Discussion

The importance of B cells in RRMS pathogenesis has become clear through many recent studies. Here we demonstrate several important differences in subset composition and phenotype between RRMS patients and healthy controls. RRMS patients had a significant increase in the frequency of B cells in the PBMC fraction. Examining B cell subsets, RRMS patients had fewer memory cells and more B1 cells in circulation when compared with healthy controls, the memory cells showed a higher proportion of the IgD-switched memory phenotype and the B1 cells showed a higher proportion of CD5 expression (an activation marker). In some or all B cell subsets, RRMS patients had a lower frequency of CD95 expression, higher PD-1, higher PD-L1, lower Siglec-10, lower CD22, higher CD305 and higher CD307d. Expression levels as measured by MFI showed similar differences as well.

There are multiple mechanisms to achieve homeostasis after an inflammatory response. One mechanism by which the immune system controls immune responses is through elimination of activated cells through apoptosis. A primary means by which lymphocytes are driven to undergo apoptosis is through Fas/FasL interactions. Fas is upregulated on activated lymphocytes, including B cells, and when bound by FasL causes the cells to form the death-inducing signaling complex, which leads to apoptosis. Fas/FasL interactions are critical to maintaining immune homeostasis, and the lack of Fas leads to autoimmune disease in both mice and humans. In EAE, Fas-driven apoptosis of B cells has been shown in the CNS during the recovery phase of the disease ^28^. Patients with MS show high levels of Fas/FasL in brain lesions. We observed that B cells of all subsets we examined (naïve, memory and B1) had a lower frequency of expression of Fas when compared with healthy controls, and that the B1 subset in healthy individuals expressed at least double the level of expression of Fas compared to patients. Interestingly, the B1 subset itself was significantly increased in the blood of RRMS patients. These results indicate that this apoptosis-inducing pathway may be defective in patients with RRMS, and potentially indicates a means by which pathogenic B cells escape homeostatic control by the immune system in this disease.

Another means by which immune responses can be controlled is through direct inhibition of activation. While its effects on T cells have been well studied, recently PD-1 on human B cells was shown to associate with the BCR and decrease activation ^29^. We observed a significant increase in frequency PD-1 expression in naïve and memory B cells in patients with RRMS, with a concomitant increase in expression in the PD-1 expressing B cells. The frequency on the B1 subset was similar to healthy controls, but the expression was significantly higher. These results may indicate a compensatory mechanism by which immune activation is controlled in these individuals due to the lower frequency of Fas expression in these B cells. The ligands for PD-1, PD-L1 and PD-L2 show different patterns of expression, with PD-L1 being widely expressed on hematopoietic and non-hematopoietic cells and PD-L2 generally being restricted to APCs. We observed high frequency of PD-L1 expression on all subsets of B cells examined, with small but significant increases seen in RRMS patient memory and B1 B cells. PD-L2 showed similar frequencies of expression on all subsets, but small but significant increases in expression levels in all three B cell cell subsets in RRMS patients. These increases in ligand expression may indicate an attempt by the immune system to decrease activation in PD-1 expressing T cells and B cells in light of the increased activation that is seen with disease.

Proteins containing ITIMs are critical modulators of the immune response. They typically increase with activation and activate phosphatases that act upon phosphorylated activation-induced signal transduction molecules which can serve to dampen immune responses. The siglecs are a family of sialic acid binding transmembrane proteins that contain ITIM domains and serve to dampen immune activation. Two prominent siglecs involved in B cell function are CD22 and siglec-10. We observed lower frequencies of expression of both of these proteins in RRMS patients. Siglec-10 showed a lower frequency of expression on naïve, memory and B1 B cells, which may indicate an overall dysfunction in siglec-10 mediated regulation of B cell activation. However, in the naïve and B1 cell subsets, the Siglec-10 positive cells that were present had a higher relative expression of the protein when compared with healthy controls, potentially showing that this smaller subset of cells may be more susceptible to regulation than those from healthy controls. CD22 showed a higher frequency of expression overall compared to siglec-10, and RRMS patients showed a slight decrease in frequency in naïve B cells and a more substantial decrease in the B1 B cell population. In contrast to what was seen with siglec-10, cell surface expression of CD22 was decreased in all three cell subsets. These observations of CD22 indicate another potential defect in the ability of B cells from RRMS patients to be downregulated.

CD305 (LAIR-1) and CD307d (FcRL4) are Ig superfamily members containing ITIMs found on the surface of some B cells. CD305 or LAIR-1 is a collagen receptor that has also been shown to bind the complement component C1q to inhibit dendritic cell activation. Other reports have focused on the ability of LAIR-1 to control type I interferon expression by dendritic cells. In the present study, we observed a high frequency of CD305 expression on naïve B cells and B1 B cells, with a slightly reduced frequency on memory B cells. This frequency was slightly increased in RRMS patients on both memory and B1 B cells. Surface expression on the CD305+ B1 B cells was also increased in RRMS patients. CD307d or FcRL4 frequency was much lower overall, but was increased in both the naïve and memory B cell populations in RRMS patients. Conversely, the level of expression on the CD307d+ cells in RRMS patients was reduced in both the memory and B1 B cell subsets. Taken together, the expression patterns of these ITIM-containing Ig superfamily members indicates cells in RRMS patients that may be slightly more susceptible to inhibition of B cell activation through ligation of these receptors.

In this study we demonstrate significant differences in B cell subset composition and phenotype in RRMS patients, especially in relation to markers that may play an important role in immune regulation and homeostasis. The balance of changes seem to indicate B cells that are more resistant to common means of regulation (CD95 and siglec-mediated) and perhaps less resistant to alternative means of regulation (PD-1 and ITIM-containing Ig superfamily-mediated). This unique phenotypic signature seen in RRMS patients may provide a means for screening for disease or assessing B cell-directed or other therapies. Finally, these markers may provide new targets for B cell-mediated therapy of MS that could be more specific than current B cell ablating approaches.

## Data Availability

All data produced in the present study are available upon reasonable request to the authors.

## Acknowledgements

This work was supported by funding from Bayer Healthcare Pharmeceuticals. Studies were performed in DartLab, the Immunoassay and Flow Cytometry Shared Resource at the Geisel School of Medicine, which receives support from the Norris Cotton Cancer Center and the Dartmouth COBRE Center for Molecular, Cellular and Translational Immunological Research.

