## Supplementary material for "Circulating B Cells in Relapsing-Remitting Multiple Sclerosis Show Markedly Different Patterns of Regulatory Marker Expression Compared with Healthy Controls": Table 1

**Table 1. Antibodies used for B Cell Surface Staining Panels**

| **Epitope** | **Fluorochrome** | **Clone** | **Supplier** | **Panel** |
| --- | --- | --- | --- | --- |
| CD43 | FITC | 1G10 | BD | All |
| CD27 | PE-CF594 | MT271 | BD | All |
| CD24 | PerCP-eFluor® 710 | SN3 A5-2H10 | eBioscience | ITIM, |
| CD19 | PE-Cy7 | H1B19 | Biolegend | All |
| CD20 | Alexa Fluor 700 | 2H7 | Biolegend | All |
| CD69 | APC-Cy7 | FN50 | Biolegend | All |
| CD5 | PerCP-Cy5.5 | UCHT2 | Biolegend | PD |
| CD14 | VioGreen | TUK4 | Miltenyi | All |
| IgD | PE | IA6-2 | Biolegend | Subset |
| CD95 | BrV421 | DX2 | Biolegend | ITIM |
| PD-1 | PE | EH12.2H7 | Biolegend | PD |
| PD-L1 | BrV421 | 29E.2A3 | Biolegend | PD |
| PD-L2 | AF647 | 24F.10C12 | Biolegend | PD |
| SIGLEC-10 | PE | 5G6 | Biolegend | ITIM |
| CD22 | APC | HIB22 | Biolegend | ITIM |
| CD305 | PerCP-Cy5.5 | NKTA255 | Biolegend | Costim |
| CD307d | PE-Cy7 | 413D12 | Biolegend | ITIM |
| IgM | APC | MHM-88 | Biolegend | Costim |
| CD80 | BrV421 | 2D10 | Biolegend | Costim |
| CD152 | PE | L3D10 | Biolegend | Costim |
| CD86 | PE-Cy7 | IT2.2 | Biolegend | Costim |
